# Associations Between Plasma Omega-3, Fish Oil Use and Risk of AF in the UK Biobank

**DOI:** 10.1101/2025.04.04.25325263

**Authors:** Evan O’Keefe, James H O’Keefe, Nathan L Tintle, W Grant Franco, Jason Westra, William S Harris

## Abstract

**Objective:** To determine the relationship between plasma omega-3 levels and incident atrial fibrillation (AF), and the association between fish oil supplement (FOS) use and risk for AF.

**Methods and Analysis:** Recent studies in UK Biobank concluded that FOS use was associated with increased risk of incident AF. Conversely, a meta-analysis found inverse relationships between blood levels of omega-3 and AF risk. We performed a prospective observational study linking plasma omega-3 levels and reported FOS use with AF risk in UK Biobank. Among UK Biobank participants without prevalent AF, 261 108 had plasma omega-3 levels and 466 169 reported FOS use. The primary outcome was incident AF during follow-up (median 12.7 years). Multivariable-adjusted hazard ratios (HR, 95% confidence intervals, CI) for fatty acids were computed continuously (per inter-quintile range, IQ_5_R) and by quintile. Hazard ratios were computed for dichotomous fish oil supplement use.

**Results:** Plasma omega-3 levels were inversely associated with incident AF (HR per IQ_5_R = 0.90, 95% CI 0.86, 0.93; HR=0.87 [0.83, 0.91] in quintile 5 vs quintile 1). Fish oil supplement use was reported by 31% of the cohort and was more common in older individuals. After adjusting for age as a continuous variable, no association was observed between fish oil supplement use and AF risk (HR=1.00; 95% CI 0.97, 1.02).

**Conclusion:** Higher circulating omega-3 levels were linked to reduced AF risk in UK Biobank. Further, after age was adjusted for as a continuous variable, no association was found between fish oil supplement use and AF.

## INTRODUCTION

The most prevalent sustained cardiac arrhythmia is AF (AF), with > 3 million new cases each year and a global prevalence of >37 million people. Moreover, it is a significant public health concern, elevating risks of hospitalization, premature mortality, heart failure, and thromboembolic events including stroke.^1^ Higher blood levels of omega-3 fatty acids have been favorably associated with lower risk for all these adverse outcomes, however their association with AF has been less clear.

Randomized trials assessing pharmaceutical omega-3 products containing EPA and/or DHA have suggested a dose-dependent increase in risk of AF at doses ranging from 1 to 4 g/day.^2^ ^3^ However, contrasting data from a meta-analysis of 17 biomarker-based observational studies, nearly 55,000 individuals in total, found that increasing blood levels of omega-3 were associated with lower risk of AF.^4^

Using UK Biobank data, one research group published two recent reports concluding that self-reported use of fish oil supplements (FOS, over-the-counter sources of EPA and DHA) was associated with an increased risk of incident AF.^5^ ^6^ This finding is at odds with previous studies demonstrating improved cardiovascular outcomes and a reduced risk of AF associated with higher plasma levels of omega-3s.^4^ In light of these conflicting findings, we sought to further investigate the relationship between omega-3 levels and AF risk in the UK Biobank using serum omega-3 levels as the exposure, and to re-evaluate the link between self-reported FOS use and AF.

## MATERIALS AND METHODS

### Sample

The UK Biobank is a prospective, population-based cohort of 502 366 individuals, aged 40-69 years, recruited in the UK between April 2007 and December 2010.^7^ UK Biobank has ethical approval (Ref. 11/NW/0382) from the Northwest Multi-centre Research Ethics Committee as a Research Tissue Bank. All participants gave electronic signed informed consent. The UK Biobank study was conducted according to the guidelines established by the Declaration of Helsinki. The UK Biobank protocol is available online (http://www.ukbiobank.ac.uk/wp114content/uploads/2011/11/UK-Biobank-Protocol.pdf). The University of South Dakota Institutional Review Board approved the use of these de-identified, publicly available data for research purposes (IRB-21-147). Within the cohort, 274 123 had data on plasma fatty acids. Of these, 4639 had prevalent AF and another 8376 were missing at least one covariate, leaving 261 108 as the primary analytic sample size. Secondary analyses used a larger sample of individuals (n=466 169) who answered a question on self-reported FOS use, did not have prevalent AF, and were not missing continuous covariates, regardless of whether blood fatty acid data were available.

### Outcome Assessment

Date of first diagnosis of AF is available via the primary UK Biobank dataset (variable source code: 131350), and is primarily based on electronic medical record information from hospital admissions or primary care, with a limited number of events based on death registries or self-report (source of AF available in UK Biobank: https://biobank.ndph.ox.ac.uk/showcase/field.cgi?id=131351). Individuals with date of first AF before measurement of plasma fatty acids were dropped from the analytic sample (see previous paragraph). A complete list of UK Biobank variable identification corresponding to covariates, exposures and outcomes is available in **Supplemental Tables 1 and 2**.

### Exposure Assessment

The UK Biobank includes data on two plasma omega-3 metrics: DHA and total omega-3s. We constructed a third metric, other omega-3 fatty acids (i.e., the sum of alpha linolenic acid, EPA, and docosapentaenoic acid), as the difference between these two. Plasma samples were collected at baseline and analyzed for omega-3s by nuclear magnetic resonance (Nightingale Health Plc, Helsinki, Finland).^8^ We note that in a recent inter-laboratory experiment ^9^ comparing fatty acid determinations by nuclear magnetic resonance and gas chromatography, the strongest predictor of other omega-3 % was EPA % (R^2^=47%). Equations to convert plasma omega-3 metrics measured by nuclear magnetic resonance to plasma and red blood cell metrics measured by gas chromatography, based on Schuchardt et al.^9^ are provided in **Supplemental Table 3**. Omega-3 biomarkers from the nuclear magnetic resonance analysis were analyzed as individual quintiles or after standardization by the inter-quintile range of the fatty acid (e.g., dividing each fatty acid metric by the difference between the 90^th^ and 10^th^ percentile). In separate analyses we considered self-reported FOS use (Yes/No; UK Biobank Variable Identification: 6179) as a fourth exposure.

### Covariate Assessment

In our multivariable models for the analysis of risk for AF by plasma omega-3 levels, we adjusted for the following demographics, behavioral, biomarker- and health-related variables: biological sex (male/female), body mass index, self-reported race/ethnicity (Asian, Black, White, Other), education (college, high school, less than high school), self-reported alcohol consumption (rarely, monthly, 1-2x/week, 3-4x/week, daily), self-reported exercise (quartiles of moderate-to-vigorous exercise; as minutes/week), smoking status (never, previous, current), plasma linoleic acid, and other plasma omega-6 fatty acids (total omega-6 fatty acids minus linoleic acid), self-reported use of beta-blockers, self-reported treatment for hypertension, self-reported treatment for high cholesterol, prevalent diagnosis of diabetes, prevalent diagnosis of cardiovascular disease, prevalent diagnosis of heart failure.

Covariates used in our replication the FOS and risk for AF report by Zhang et al.^6^ were those used by Zhang and are listed in **Supplementary Table 4**. These were similar to those listed above with the exception that variables like oily fish intake, the Townsend deprivation index, and a genetic risk score were included, and naturally, plasma omega-6 fatty acid levels were not (since they were not available on the full cohort used in this analysis).

### Statistical Analyses

Descriptive sample characteristics were summarized using standard statistical methods. Cox-proportional hazards models were used to estimate adjusted hazard ratio and associated 95% CIs between incident AF (time to event or censoring) with DHA %, total omega-3 % or other omega-3 %, per quintile using quintile 1 as reference, as well as (in separate models), inter-quintile range, and using covariates as noted above. Tests of non-linearity for omega-3 fatty acid biomarkers were conducted using cubic splines. Fixed-effects meta-analysis was used to combine UK Biobank results with a recently conducted meta-analysis.^4^ In parallel models examining the association between self-reported FOS use (yes/no) and incident AF, we considered different ways of adjusting for age (1) continuously (linear), (2) continuously (cubic splines), (3) dichotomously (65+ vs. <65) or (4) categorically (<45, 45-49, 50-54, 55-59, 60-64, 65+). Sensitivity analyses explored the use of different covariate sets and different dates for data censoring (in order to replicate the prior studies on this topic). Statistical significance was set to 0.05 for all analyses, except for tests of non-linearity which used 0.01 due to the exploratory and multiple-testing nature of those tests, with two-sided tests in all cases. Proportional hazards assumptions were confirmed. R (www.r-project.org) was used for all analyses including the coxme and metafor packages.

## RESULTS

### Associations of plasma omega-3 fatty acids with incident AF

Amongst the 261 108 individuals with plasma fatty acid data available in the UK Biobank, the mean age was 56.5 ± 8.1 years, 95% were white and 55% were female with an average body mass index of 27.4 ± 4.8 (kg/m^2^) (**Table 1**). After a median follow-up of 12.7 years, all three omega-3 fatty acid biomarkers showed statistically significant, inverse associations with incident AF risk (**Table 2**). When comparing the highest to lowest quintiles for total omega-3, risk for AF was 14% lower; for other omega-3s, the adjusted risk was 16% lower, and for DHA, 8% lower. Absolute risk for AF in Q1 total omega-3 was 7.5% and 6.9% in Q5. Similar inverse associations were seen in the per inter quintile range analyses. There was no evidence of non-linearity in the biomarker relationships (p>0.01 in all cases).

**Table 1.** Subject Characteristics (N=261 108)

| **Variable** | **% (count) or Mean (SD)** |
| --- | --- |
| Age (years) | 56.5 (8.1) |
| Sex – Female | 54.6% (142 617) |
| Race/Ethnicity |  |
| White | 95.4% (249 021) |
| Asian | 1.9% (5013) |
| Black | 1.4% (3543) |
| Other | 1.4% (3531) |
| BMI (kg/m<sup>2</sup>) | 27.4 (4.8) |
| Education |  |
| College | 60.2% (157 223) |
| High School | 22.5% (58 737) |
| Less than High School | 17.3% (45 148) |
| Exercise |  |
| Lowest Quartile | 22.3% (58 329) |
| Second lowest Quartile | 23.0% (59 970) |
| Second highest Quartile | 23.1% (60 344) |
| Highest quartile | 23.3% (60 914) |
| Unavailable | 8.3% (21 551) |
| Smoking status |  |
| Never | 54.8% (143 156) |
| Previous | 34.7% (90 516) |
| Current | 10.5% (27 436) |
| Alcohol consumption |  |
| Rarely | 30.2% (78 733) |
| 1-2x/week | 26.2% (68 446) |
| 3-4x/week | 23.4% (61 139) |
| Daily | 20.2% (52 790) |
| Linoleic Acid (% composition) | 28.9 (3.4) |
| Non-Linoleic Omega-6s (% composition) | 8.9 (1.9) |
| Taking Beta-Blockers | 6.0% (15 637) |
| Treated for Hypertension | 20.4% (53 368) |
| Treated for high cholesterol | 17.2% (44 966) |
| Diagnosed with Diabetes | 5.1% (13 317) |
| Diagnosed with Cardiovascular Disease | 6.3% (16 469) |
| Diagnosed with HF | 0.4% (995) |

**Table 2.**
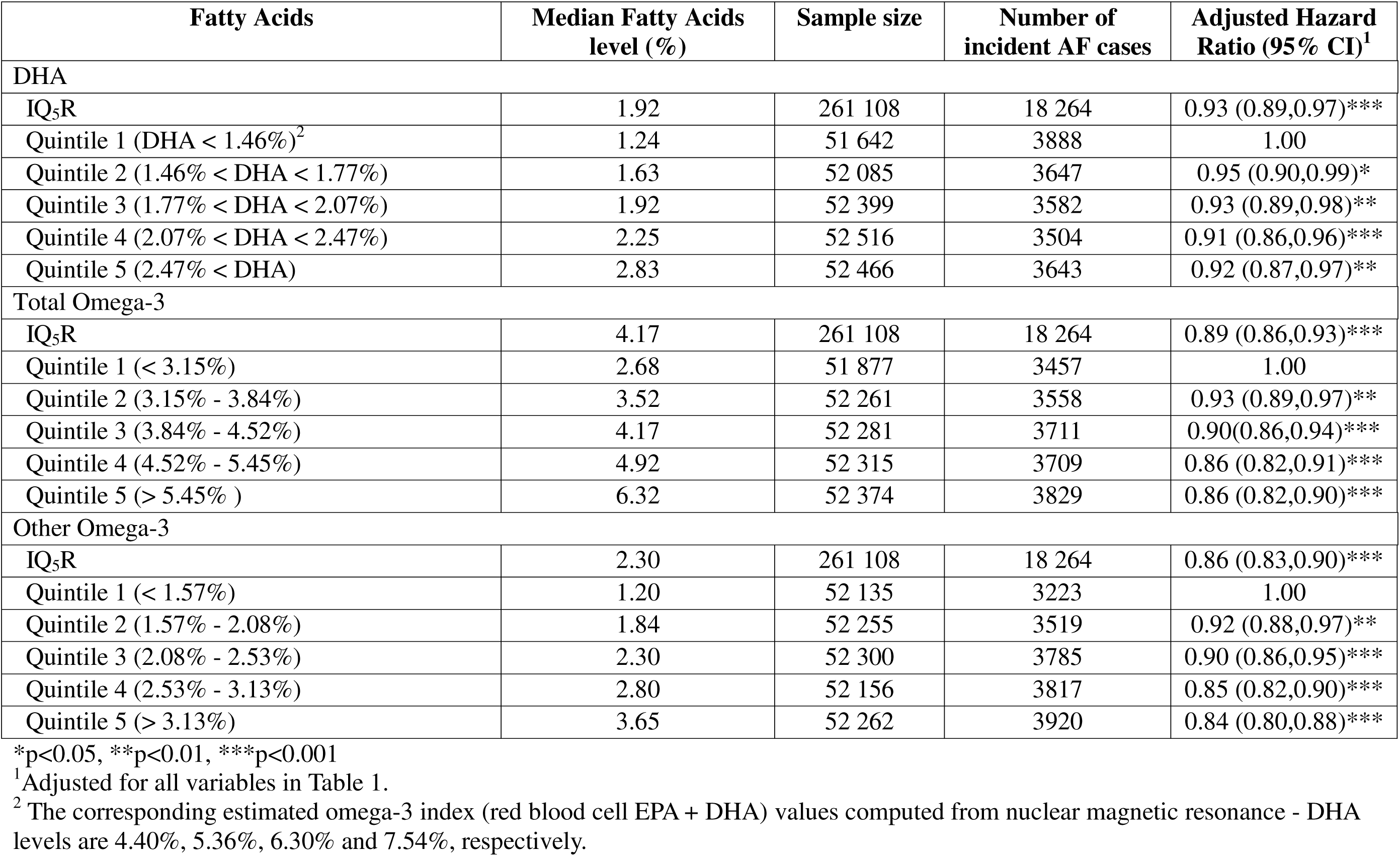
Association of Plasma Omega-3 Quintiles with Prospective AF in the UK Biobank.

### Update of Qian et al. Meta-analysis with UK Biobank

Qian et al. analyzed 17 prospective cohort studies, encompassing 55,214 individuals among whom there were 7720 incident cases of AF during a median follow-up of 13.3 years.^4^ Since the current analysis used the same covariate set as Qian et al., we were able to harmonize and update the meta-analysis with the present results from the UK Biobank (**Table 3**) now including 316 322 persons. Originally, Qian et al. found inverse relationships between DHA, DHA and EPA + DHA and risk of AF. The findings were largely similar in the updated meta-analysis in which we used the equations in **Supplemental Table 3** to predict plasma docosapentaenoic acid, EPA and EPA + DHA levels from the UK Biobank nuclear magnetic resonance data. The only difference was that for EPA, Qian et al. found no association between blood EPA and incident AF, while the UK Biobank analysis showed a significant inverse association. The inclusion of UK Biobank data did not substantially change the observed heterogeneities in the meta-analysis (52.2% to 59.7%).

**Table 3.** Meta-analysis from Qian et al.^1^ updated with UK Biobank Findings.

| Fatty acid | Analysis | Adjusted Hazard Ratio per IQ<sub>5</sub>R<sup>4</sup> | Heterogeneity I<sup>2</sup> (p-value) |
| --- | --- | --- | --- |
| DHA | Original meta-analysis<sup>1</sup> | 0.90 (0.85, 0.96) ** | 47.5% (p=0.016) |
|  | UK Biobank alone | 0.93 (0.89, 0.97) *** | -- |
|  | Updated meta-analysis<sup>2</sup> | 0.92 (0.89, 0.96) *** | 43.4% (p=0.026) |
| EPA<sup>3</sup> | Original meta-analysis<sup>1</sup> | 1.00 (0.95, 1.05) | 52.2% (p=0.008) |
|  | UK Biobank alone | 0.92 (0.88, 0.95) *** | -- |
|  | Updated meta-analysis<sup>2</sup> | 0.94 (0.92, 0.97) *** | 59.7% (p=0.0009) |
| Docosapentaenoic acid<sup>3</sup> | Original meta-analysis<sup>1</sup> | 0.89 (0.83, 0.95) *** | 0.0% (p=0.51) |
|  | UK Biobank alone | 0.91 (0.87, 0.94) *** | -- |
|  | Updated meta-analysis<sup>2</sup> | 0.90 (0.88, 0.93) *** | 0.0% (p=0.56) |
| EPA + DHA<sup>3</sup> | Original meta-analysis<sup>1</sup> | 0.93 (0.87, 0.99) * | 60.7% (p=0.001) |
|  | UK Biobank alone | 0.92 (0.88, 0.95) *** | -- |
|  | Updated meta-analysis<sup>2</sup> | 0.92 (0.90, 0.95) *** | 58.5% (p=0.0013) |

\*p<0.05, \*\*p<0.01, \*\*\*p<0.001

### Re-evaluation of Prior Findings on Fish Oil Supplement Use and Risk of AF in the UK Biobank

As noted, two recent papers on risk for incident AF and self-reported FOS use in UK Biobank reported increased risk among consumers of FOS.^5^ ^6^ The 2022 and the 2024 studies reported identical AF hazard ratios and confidence intervals among individuals without prior cardiovascular disease (adjusted hazard ratio 1.13; 95% CI: 1.10, 1.17). We were able to replicate these findings if age was adjusted for as a *dichotomous* variable (e.g. age 65+ *vs* <65) (**Table 4**). However, when age was treated as a *continuous* variable the associations between FOS use and incident AF was lost (adjusted hazard ratio = 1.00; 0.97, 1.02). Other approaches of adjusting for age-related risk (splines, more age categories), yielded findings similar to those noted when age was adjusted for as a continuous variable (**Table 4**). In addition, models adjusting for age in a more continuous manner showed improved concordance (i.e., predictive ability) compared with those using dichotomous age (p<0.0001), providing additional confirmation of the superiority of more granular age adjustments **(Table 4)**. This approach was further confirmed in our sensitivity analyses (**Table 4**), which considered different follow-up periods and different covariate sets (see **Supplemental Table 4** for distributions of these variables).

**Table 4.** Re-evaluation of the association between fish oil supplement use and risk for AF in Zhang et al.

|  | Sample size | Fish oil users | Total # Incident AF | Type of age adjustment | Adjusted Hazard Ratio <sup>1</sup> | Adjusted Hazard Ratio <sup>2</sup> | Model Concordance |
| --- | --- | --- | --- | --- | --- | --- | --- |
| Previously published results<sup>3</sup> | 468 665 | 148 192 | 25 748 | None/ Dichotomous | 1.22 (1.19, 1.25)<sup>4</sup> | 1.10 (1.07, 1.13)<sup>5</sup> |  |
| Replication attempts – data censored at 9/1/2020<sup>5</sup> | 454 568 | 143 736 | 26 667 | None | 1.20 (1.17,1.23) | -- |  |
|  |  |  |  | Dichotomous | 1.05 (1.02,1.08) | 1.09 (1.06,1.12) | 0.746 |
|  |  |  |  | Continuous – linear | 0.92 (0.90,0.95) | 0.99 (0.97,1.02) | 0.773 |
|  |  |  |  | Continuous – Spline | 0.92 (0.90,0.95) | 0.99 (0.97,1.02) | 0.773 |
|  |  |  |  | 5-year age category | 0.93 (0.91,0.95) | 1.00 (0.97,1.02) | 0.770 |
| Replication attempts – data censored at 12/31/2022<sup>6</sup> | 454 568 | 143 736 | 32 162 | No Age Adjustment | 1.21 (1.18, 1.24) |  |  |
|  |  |  |  | Dichotomous | 1.06 (1.03,1.08) | 1.10 (1.07, 1.12) | 0.742 |
|  |  |  |  | Continuous – linear | 0.93 (0.91, 0.95) | 1.00 (0.97, 1.02) | 0.770 |
|  |  |  |  | Continuous - Spline | 0.93 (0.91, 0.95) | 1.00 (0.98, 1.02) | 0.770 |
|  |  |  |  | 5-year age category | 0.94 (0.92,0.96) | 1.00 (0.98,1.03) | 0.768 |

## Discussion

We found that higher circulating levels of omega-3s were associated with reduced risk for AF in the UK Biobank. Fully adjusted risk reductions when comparing the highest to lowest quintiles for total omega-3, other omgea-3, and DHA were 14%, 16%, and 8% respectively. These findings largely paralleled those from a recent meta-analysis by Qian et al. pooling data from 17 cohorts and including nearly 55,000 individuals who demonstrated a 12% decreased risk of AF for patients in the top quintile of DHA + EPA blood levels compared to the bottom quintile.^4^ After the present data were added into the aforementioned meta-analysis, higher blood levels of DHA, EPA, docosapentaenoic acid and EPA + DHA were associated with a 6 to 10% lower risk of AF. Finally, and importantly, a re-analysis of the relationship between risk of AF and the use of FOS in the UK Biobank revealed that, after adjusting for age as a continuous variable, the previously reported adverse association between the two disappeared.

Previous meta-analyses of observational studies that have focused on biomarkers instead of estimated intakes have shown that higher levels of marine omega-3s, which are reflective of chronic dietary intake, correlate with lower risks for all-cause mortality and cardiovascular mortality,^10^ ^11^ stroke,^12^ and most recently, AF.^4^ The Qian study, along with the findings presented here, sharply contrast with two previous reports,^5^ ^6^ which were both from the same research group, both used the same UK Biobank database, and both concluded that FOS use may increase the risk of AF. Since FOS use is directly related to serum omega-3 polyunsaturated fatty acid levels,^9^ the findings of these two previous studies are at odds with our observations. To explore this apparent contradiction, we undertook a reanalysis of the same UKBB data used by Zhang et al.^6^ and Chen et al.^5^ In both of these studies, age was adjusted for as a dichotomous, not a continuous, variable. Since the incidence of AF and the use of FOSs both increase linearly with age in the UK Biobank (**Figure 1**), a simple dichotomous adjustment for age is insufficient. When age was treated as a *continuous* variable the apparent association between FOS use and AF disappeared.

**Fig 1.**
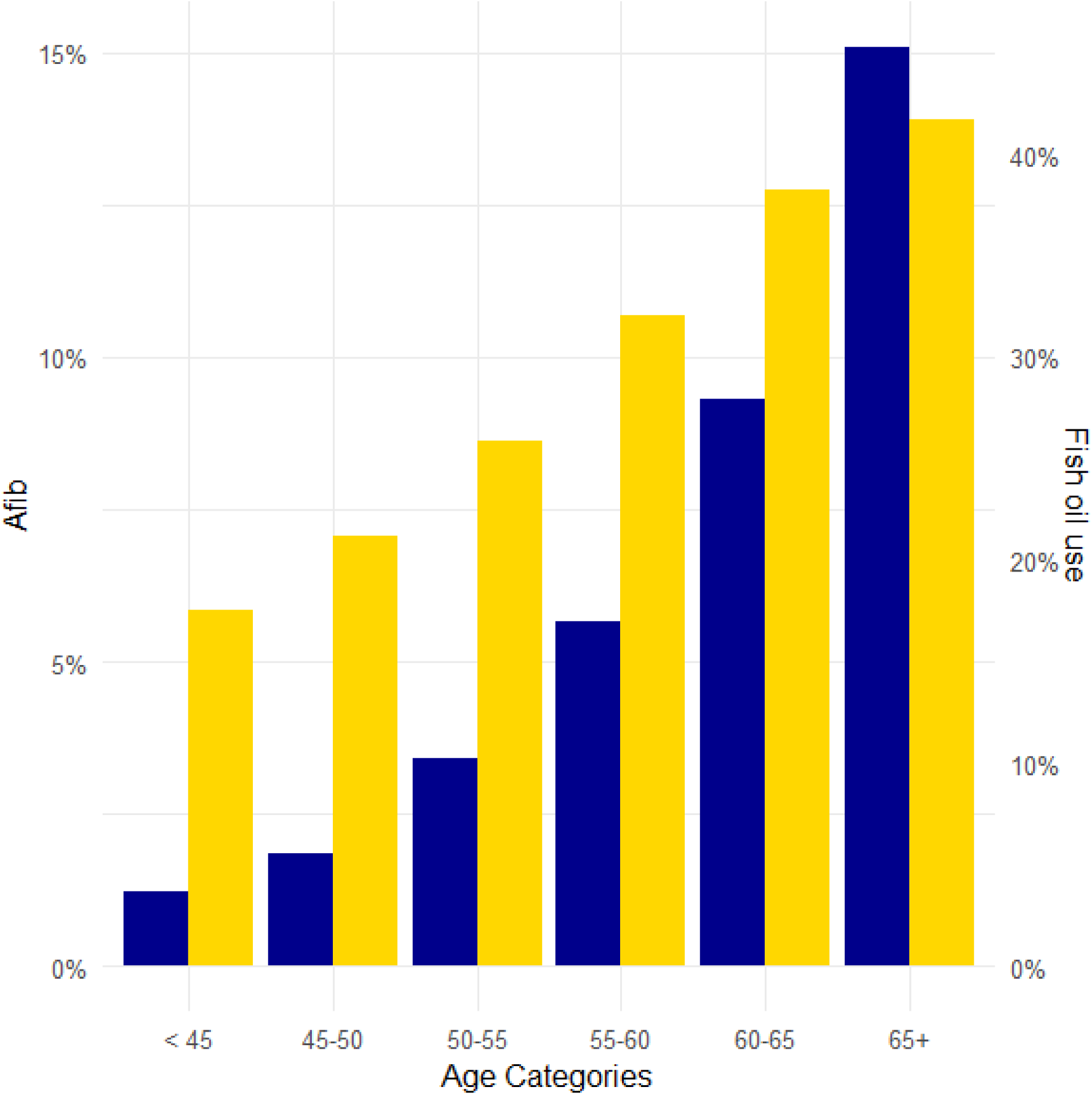
Reported use of Fish Oil Supplements at Baseline and Incidence of AF by Age at Baseline in the UK Biobank. The parallel age-related increases in fish oil use (yellow) and incident AF (blue) require rigorous statistical age adjustment.

In a recent meta-analysis of randomized controlled trials testing the effects of pharmaceutical omega-3 products in 83,112 individuals, there was a 24% increase in relative risk of AF (p=0.0002).^3^ This relationship appeared to be dose-dependent with a 12% higher relative risk of AF in the five trials testing <1000 mg/d of DHA + EPA, versus a 51% relative risk increase of AF in the three trials that used 1.8 to 4.0 g/d of DHA and/or EPA. The weighted absolute increase in risk of AF in these studies were 0.67% overall, 0.46% for lower dose omega-3 trials, and 1.18% for the high dose trials (**Supplemental Table 5** including a new report^13^ not included in Jia et al.^3^ meta-analysis). These relative risk increases have been questioned, however, by Samuel and Nattel who noted that since the risk for dying in all but one of these trials was three to five times higher than the risk for developing AF, and because the individuals randomized to omega-3 had a significantly lower risk of dying compared to placebo-treated patients, a failure to adjust for competing risks may have inflated the relative risk estimates.^14^

These apparently contradictory findings regarding AF risk between randomized trials and biomarker- and FOS use-based observational studies is a currently unresolved issue. Adding to the confusion is the fact that 20 years ago, there was sufficient evidence to mount a major trial in 663 patients with AF in which high dose omega-3 (4 to 8 g/d) were given to *reduce* recurrent AF episodes.^15^ The hypothesis was not supported by the data (indeed, there was a trend towards *higher* risk for recurrence in the omega-3 group), but this underscores why there remains significant uncertainty about the role of omega-3 and AF. New randomized controlled trials designed to address this important question are needed.

Mechanistically, a variety of possibilities have been suggested as to why higher omega-3 fatty acid levels would be associated with susceptibility to AF. As omega-3 fatty acids become incorporated into myocardial membranes, they may alter the conformation of PIEZO1 and PIEZO2 channels.^16^ These membrane-bound pressure-sensing proteins may be activated when high levels of omega-3s are in the membranes and become arrhythmic triggers. On the other hand, low levels of omega-3s can predispose to heart failure, hypertension, atherosclerotic cardiovascular disease and other cardiac conditions that increase risk for AF^17^. Intermediate omega-3 levels, such as those found in individuals consuming fish/seafood and/or low to moderate doses of over-the-counter fish oil, but not high-dose pharmaceutical products, may be protective, resulting in a potential U-shaped relationship between omega-3 status and AF risk (**Figure 2**).

**Fig 2.**
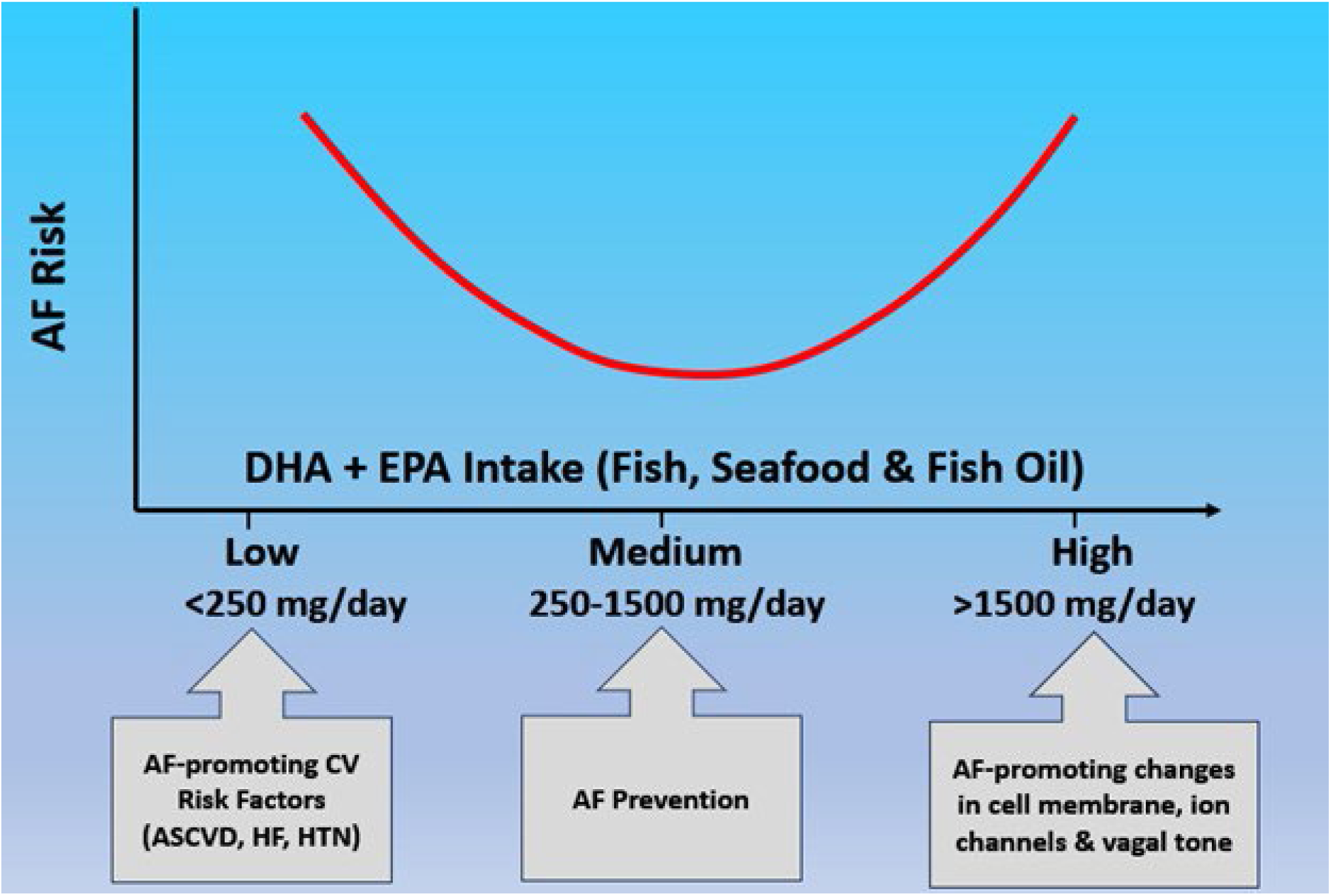
U-shaped Relationship Between DHA + EPA Intake & Risk of AF. The chronic risk of AF appears to be higher for individuals consuming low amounts of EPA + DHA (<250 mg/day) and large amounts (>1500 mg/day). The AF risk is lowest for those consuming moderate amounts of EPA + DHA (250 to 1500 mg/day). Figure adapted from Fatkin et.al.^16^

Other possible explanations relate to vagal tone.^18^ Studies have consistently shown that omega-3s produce a dose-dependent increase in vagal tone which becomes apparent starting at relatively low doses of EPA + DHA (∼500 mg/d).^19–21^ Low-level vagal nerve stimulation is antiarrhythmic and is associated with a reduced risk of AF.^22^ In contrast, high-level vagal nerve stimulation is an effective way to induce AF in animal models.^23^ This phenomenon suggests another possible explanation of how lower-dose (dietary) omega-3 intakes correlate with reduced risk of AF, whereas high-dose (pharmaceutical) omega-3 use appears to be associated with increased risk of AF.

Prior dietary questionnaire-based epidemiological studies have suggested that an intake of ∼500 to 700 mg/d of DHA + EPA may be the ideal dose for decreasing risk of AF.^24^ A large observational study found that consumption levels less than this were associated with ∼10% higher risks of AF,^24^ whereas omega-3 intakes above 1,250 mg/d are also linked with increased risks of AF.^25^ The typical adult American eats <1 serving/week of fish or seafood; so the average intake of DHA + EPA in the United States is approximately 100 mg/d.^26^ ^27^ This corresponds to an average DHA + EPA level in red blood cells (i.e., the omega-3 index) in the United States of about 5.4%.^28^ The estimated median omega-3 index in quintile 5 in the present study was 8.62%. The omega-3 index level that is optimal for minimizing risk of major adverse cardiovascular events, stroke and all-cause mortality is ≥8%; to achieve this would require an increase of approximately 1500 mg/d of DHA + EPA.^11^ ^26^

However, for people at high risk for AF or those with a history of AF, a lower target of ∼500 to 700 mg/d of DHA + EPA, preferably from fish/seafood, would seem to be a safer level of intake.^29^ The 2021 American Heart Association dietary guidance for improving cardiovascular health advocates for a diet that prioritizes the consumption of vegetables, fruits, and fish/seafood as a preferred source of protein. The American Heart Association also specifically recommends at least two meals per week that feature fish or seafood, ^30^ which in the case of salmon would supply ∼300 mg of EPA + DHA per day.

Finally, it is of interest to note that the omega-3 index in Japan and South Korea averages >8% compared with about 5.5% in the United States,^31^ yet the age-adjusted prevalence of AF in 2019 in the former two countries was 200-399 compared with >1200 per 100 000 in the United States. This ecological finding is at least consistent with the results of the current observational study— higher intakes (thus blood levels) of omega-3 are associated with lower risks of AF, although obviously causal inferences cannot be drawn.

### Strengths and Limitations

One strength of this study was the use of objective measurements of omega-3 plasma levels rather than relying on dietary questionnaires. Furthermore, the large sample of > 261 000 participants followed for 12 years gives us sufficient statistical power to detect relationships between omega-3 and AF. A limitation of the UK Biobank is the relative lack of ethnic diversity as nearly all individuals are White. However, the UK Biobank data harmonized strongly with a recent meta-analysis of 17 international cohorts evaluating omega-3 blood levels and risk of AF,^4^ thus are findings are likely to be generalizable. DHA and other omega-3 fatty acids and all covariates were measured only at baseline; however, omega-3 blood levels have shown good reproducibility over time.^32^ The UK Biobank cohort is generally healthier than the UK population, however, the biomarker–disease associations observed in the UK Biobank cohort are considered to be generalizable.^33^ Because this is an observational study, causation cannot be established. Even though we made statistical adjustments for many relevant risk factors (age, sex, occupation, education, physical activity, smoking, hyperlipidemia, hypertension, etc.), residual confounding from other unmeasured variables is always a possibility.

## Conclusion

In agreement with recent biomarker-based meta-analyses, higher circulating blood levels of omega-3 were associated with reduced risk for AF in the UK Biobank. Secondly, this study reassessed the relationship between FOS use and risk of AF in the UK Biobank and found no evidence of an association between FOS use and AF when age is adjusted for as a continuous variable. Thus, virtually all the observational data on omega-3 dietary intake or blood levels point to higher intakes/levels reducing risk of AF.

## Data Availability

All data produced in the present study are available upon reasonable request to the authors but additional approval from the UKBiobank is necessary to utilize the original data.

## Notes

**Funding / Disclosures** O’Keefe EL – The National Heart, Lung, and Blood Institute of the National Institutes of Health under Award Number T32HL110837 supported the research reported in this publication. The content is solely the responsibility of the authors and does not necessarily represent the official views of the National Institute of Health. This project was partially supported by the Richard Galamba Foundation. JH O’Keefe is the Chief Medical Officer of Cardiotabs, a company that sells omega-3 products. WS Harris owns stock in OmegaQuant Analytics, a laboratory that offers blood fatty acid testing for researchers, clinicians, and consumers.

### Competing Interest Statement

JHO holds an interest in Cardiotabs and WSH holds an interest in Omegaquant Analytics

### Funding Statement

This project was partially supported by the Richard Galamba Foundation.

### Author Declarations

The data used in this study were obtained from the UKBiobank and are publicly available subject to the rules and policies of the UKBiobank.

## References

1. Dai H, Zhang Q, Much AA, et al. Global, regional, and national prevalence, incidence, mortality, and risk factors for atrial fibrillation, 1990-2017: results from the Global Burden of Disease Study 2017. Eur Heart J Qual Care Clin Outcomes 2021;7(6):574–82. doi: 10.1093/ehjqcco/qcaa061 [published Online First: 2020/08/01]

2. Gencer B, Djousse L, Al-Ramady OT, et al. Effect of Long-Term Marine Omega-3 Fatty Acids Supplementation on the Risk of Atrial Fibrillation in Randomized Controlled Trials of Cardiovascular Outcomes: A Systematic Review and Meta-Analysis. Circulation 2021 doi: 10.1161/circulationaha.121.055654 [published Online First: 2021/10/07]

3. Jia X, Gao F, Pickett JK, et al. Association Between Omega-3 Fatty Acid Treatment and Atrial Fibrillation in Cardiovascular Outcome Trials: A Systematic Review and Meta-Analysis. Cardiovascular drugs and therapy / sponsored by the International Society of Cardiovascular Pharmacotherapy 2021;35(4):793–800. doi: 10.1007/s10557-021-07204-z [published Online First: 2021/06/01]

4. Qian F, Tintle N, Jensen PN, et al. Omega-3 Fatty Acid Biomarkers and Incident Atrial Fibrillation. J Am Coll Cardiol 2023;82(4):336–49. doi: 10.1016/j.jacc.2023.05.024 [published Online First: 2023/07/20]

5. Chen G, Qian ZM, Zhang J, et al. Regular use of fish oil supplements and course of cardiovascular diseases: prospective cohort study. BMJ Med 2024;3(1):e000451. doi: 10.1136/bmjmed-2022-000451 [published Online First: 2024/05/27]

6. Zhang J, Cai A, Chen G, et al. Habitual fish oil supplementation and the risk of incident atrial fibrillation: findings from a large prospective longitudinal cohort study. European journal of preventive cardiology 2022;29(14):1911–20. doi: 10.1093/eurjpc/zwac192 [published Online First: 2022/09/02]

7. Sudlow C, Gallacher J, Allen N, et al. UK biobank: an open access resource for identifying the causes of a wide range of complex diseases of middle and old age. PLoS medicine 2015;12(3):e1001779. doi: 10.1371/journal.pmed.1001779 [published Online First: 2015/04/01]

8. Julkunen H, Cichońska A, Tiainen M, et al. Atlas of plasma NMR biomarkers for health and disease in 118,461 individuals from the UK Biobank. Nat Commun 2023;14(1):604. doi: 10.1038/s41467-023-36231-7 [published Online First: 2023/02/04]

9. Schuchardt JP, Tintle N, Westra J, et al. Estimation and predictors of the Omega-3 Index in the UK Biobank. The British journal of nutrition 2023;130(2):312–22. doi: 10.1017/s0007114522003282 [published Online First: 2022/10/11]

10. Harris WS, Tintle NL, Imamura F, et al. Blood n-3 fatty acid levels and total and cause-specific mortality from 17 prospective studies. Nat Commun 2021;12(1):2329. doi: 10.1038/s41467-021-22370-2 [published Online First: 2021/04/24]

11. O’Keefe EL, O’Keefe JH, Tintle NL, et al. Circulating Docosahexaenoic Acid and Risk of All-Cause and Cause-Specific Mortality. Mayo Clinic proceedings 2024;99(4):534–41. doi: 10.1016/j.mayocp.2023.11.026 [published Online First: 2024/03/20]

12. O’Keefe JH, Tintle NL, Harris WS, et al. Omega-3 Blood Levels and Stroke Risk: A Pooled and Harmonized Analysis of 1831291 Participants From 29 Prospective Studies. Stroke 2024;55(1):50–58. doi: 10.1161/strokeaha.123.044281 [published Online First: 2023/12/22]

13. Miyauchi K, Iwata H, Nishizaki Y, et al. Randomized Trial for Evaluation in Secondary Prevention Efficacy of Combination Therapy–Statin and Eicosapentaenoic Acid (RESPECT-EPA). Circulation 2024;150(6):425–34. doi: 10.1161/CIRCULATIONAHA.123.065520

14. Samuel M, Nattel S. Fish Oil Supplements May Increase the Risk for Atrial Fibrillation: What Does This Mean? Circulation 2021;144(25):1991–94. doi: 10.1161/circulationaha.121.057464 [published Online First: 2021/12/21]

15. Kowey PR, Reiffel JA, Ellenbogen KA, et al. Efficacy and safety of prescription omega-3 fatty acids for the prevention of recurrent symptomatic atrial fibrillation: a randomized controlled trial. JAMA 2010;304:2363–72.

16. Fatkin D, Cox CD, Martinac B. Fishing for Links Between Omega-3 Fatty Acids and Atrial Fibrillation. Circulation 2022;145(14):1037–39. doi: 10.1161/circulationaha.121.058596 [published Online First: 2022/04/05]

17. Tutor A, O’Keefe EL, Lavie CJ, et al. Omega-3 fatty acids in primary and secondary prevention of cardiovascular diseases. Progress in cardiovascular diseases 2024 doi: 10.1016/j.pcad.2024.03.009 [published Online First: 2024/03/29]

18. O’Keefe EL, O’Keefe JH, Abuissa H, et al. Omega-3 and Risk of atrial fibrillation: Vagally-mediated double-edged sword. Progress in cardiovascular diseases 2024 doi: 10.1016/j.pcad.2024.11.003 [published Online First: 2024/12/02]

19. Christifano DN, Chollet-Hinton L, Mathis NB, et al. DHA Supplementation During Pregnancy Enhances Maternal Vagally Mediated Cardiac Autonomic Control in Humans. The Journal of nutrition 2023;152(12):2708–15. doi: 10.1093/jn/nxac178 [published Online First: 2022/08/12]

20. O’Keefe JH, Abuissa H, Sastre A, et al. Effects of omega-3 fatty acids on resting heart rate, heart rate recovery after exercise, and heart rate variability in men with healed myocardial infarctions and depressed ejection fractions. AmJCardiol 2006;97:1127–30.

21. Tikkanen JT, Soliman EZ, Pester J, et al. A randomized clinical trial of omega-3 fatty acid and vitamin D supplementation on electrocardiographic risk profiles. Scientific reports 2023;13(1):11454. doi: 10.1038/s41598-023-38344-x

22. Stavrakis S, Stoner JA, Humphrey MB, et al. TREAT AF (Transcutaneous Electrical Vagus Nerve Stimulation to Suppress Atrial Fibrillation): A Randomized Clinical Trial. JACC Clin Electrophysiol 2020;6(3):282–91. doi: 10.1016/j.jacep.2019.11.008 [published Online First: 2020/03/21]

23. Liu L, Nattel S. Differing sympathetic and vagal effects on atrial fibrillation in dogs: role of refractoriness heterogeneity. Am J Physiol 1997;273(2 Pt 2):H805–16. doi: 10.1152/ajpheart.1997.273.2.H805 [published Online First: 1997/08/01]

24. Guardino ET, Li Y, Nguyen XM, et al. Dietary ω-3 fatty acids and the incidence of atrial fibrillation in the Million Veteran Program. Am J Clin Nutr 2023;118(2):406–11. doi: 10.1016/j.ajcnut.2023.06.001 [published Online First: 2023/06/24]

25. Rix TA, Joensen AM, Riahi S, et al. A U-shaped association between consumption of marine n-3 fatty acids and development of atrial fibrillation/atrial flutter-a Danish cohort study. Europace 2014;16(11):1554–61. doi: 10.1093/europace/euu019 [published Online First: 2014/02/28]

26. Jackson KH, Polreis JM, Tintle NL, et al. Association of reported fish intake and supplementation status with the omega-3 index. Prostaglandins, leukotrienes, and essential fatty acids 2019;142:4–10. doi: 10.1016/j.plefa.2019.01.002 [published Online First: 2019/02/19]

27. Walker RE, Jackson KH, Tintle NL, et al. Predicting the effects of supplemental EPA and DHA on the omega-3 index. Am J Clin Nutr 2019;110(4):1034–40. doi: 10.1093/ajcn/nqz161 [published Online First: 2019/08/10]

28. Schuchardt JP, Cerrato M, Ceseri M, et al. Red blood cell fatty acid patterns from 7 countries: Focus on the Omega-3 index. Prostaglandins, leukotrienes, and essential fatty acids 2022;179:102418. doi: 10.1016/j.plefa.2022.102418 [published Online First: 2022/04/03]

29. Ballantyne CM, Jia X. Omega-3 Fatty Acids and Risk for Atrial Fibrillation: Big Fish or Small Fry? J Am Coll Cardiol 2023;82(4):350–52. doi: 10.1016/j.jacc.2023.05.026 [published Online First: 2023/07/20]

30. Lichtenstein AH, Appel LJ, Vadiveloo M, et al. 2021 Dietary Guidance to Improve Cardiovascular Health: A Scientific Statement From the American Heart Association. Circulation 2021;144(23):e472-e87. doi: 10.1161/cir.0000000000001031 [published Online First: 2021/11/03]

31. Schuchardt JP, Beinhorn P, Hu XF, et al. Omega-3 world map: 2024 update. Progress in lipid research 2024:101286. doi: 10.1016/j.plipres.2024.101286 [published Online First: 2024/06/16]

32. Harris WS, Pottala JV, Vasan RS, et al. Changes in erythrocyte membrane trans and marine fatty acids between 1999 and 2006 in older Americans. The Journal of nutrition 2012;142(7):1297–303. doi: 10.3945/jn.112.158295 [published Online First: 2012/05/25]

33. Fry A, Littlejohns TJ, Sudlow C, et al. Comparison of Sociodemographic and Health-Related Characteristics of UK Biobank Participants With Those of the General Population. American journal of epidemiology 2017;186(9):1026–34. doi: 10.1093/aje/kwx246 [published Online First: 2017/06/24]

