## Supplementary Materials for "Associations Between Plasma Omega-3, Fish Oil Use and Risk of AF in the UK Biobank"

**Supplemental Table 1. Variable Definitions and Correspondence to UKBB Variable IDs.**

| **Variables** | **UKBB IDs** | **Coding of UKBB** | **Details** |
| --- | --- | --- | --- |
| AF | 131350 |  |  |
| Omega3 | 23451 |  | Percent of total fatty acids (NMR) |
| DHA | 23457 |  | Percent of total fatty acids (NMR) |
| Other Omega-3 | 23451, 23457 | 23451 - 23457 | Percent of total fatty acids (NMR) |
| LA | 23456 |  | Percent of total fatty acids (NMR) |
| Non-LA Omega6 | 23452, 23456 | 23452 - 23456 | Percent of total fatty acids (NMR) |
| Age | 21022 |  | years |
| Sex | 31 |  | Female  Male |
| Ethnicity | 21000 | White: 1001, 1002, 1003, 1  Black: 4001, 4002, 4003, 4  Asian: 3001, 3002, 3003, 3004, 5  Other: 2, 6, 2001, 2002, 2003, 2004 | White, Black, Asian, Other |
| BMI | 21001 |  | Kg/m2 |
| Cholesterol | 6153, 6177 | Yes: 1 | Yes/No |
| Smoking status | 20116 | Never: 0  Previous: 1  Current: 2 | Never, Previous, Current |
| Alcohol use | 1558 | Daily: 1  3-4x/week: 2  1-2x/week: 3  Rarely: 4, 5, 6 | Daily  3-4x/week  1-2x/week  Rarely |
| Exercise | 874/894/914  864/884/904 | Walking/Week: 874*864  Moderate Activity/Week: 894*884  Vigorous Activity/Week: 914*904  Weekly MET-like Exercise = 3.3*walking/week + 4*mod.act./week + 8*vig.act./week | Quartiles of Weekly Met-like Exercise (four categories) |
| Hypertension | 6153, 6177 | Yes: 2 | Yes/No |
| Education | 6138 | College: 1, 5, 6  High School: 2, 3, 4  Less than High School: -7 | College  High School  Less than High School |
| Diabetes | 2443 |  | Yes/No |
| Beta Blockers | 20003 | Any: 1140860192, 1140860292, 1140860308, 1140860312, 1140860316, 1140860322, 1140860332, 1140860404, 1140860418, 1140860422, 1140860426, 1140864950, 1140866724, 1140866738, 1140879760, 1140879818, 1140879824, 1140879842, 1140879854, 1140909368, 1141146124, 1141146126, 1141146128, 1141164276, 1141180778, 1141194804, 1141194810 | Yes/No |
| Prevalent MI, Stroke, PAD | 131296, 131298, 131300, 131302, 131360, 131362, 131364, 131366, 131368, 131380 | Any < Baseline | Yes/No |
| Prevalent HF | 131354, 131288,  131292 | Any < Baseline Date | Yes/No |
| Baseline Date | 53 |  |  |
| Fish Oil | 6179 | Yes: 1 | Yes/No |
| Drop Date | 191 |  |  |

**Supplemental Table 2. Covariates Used by Zhang et al.^1^**

| **Variables** | **UKBB IDs** | **Coding of UKBB** | **Details** |
| --- | --- | --- | --- |
| AF | 131350 |  |  |
| Fish Oil | 6179 | Yes: 1 | Yes/No |
| Age | 21022 |  | years |
| Age Categories | 21022 | Age < 65 | Under 65/ 65+ |
| Ethnicity | 21000 | White: 1001, 1002, 1003, 1 | White/Other |
| Sex | 31 |  | Female  Male |
| Townsend Deprivation Index | 189 |  |  |
| Oily Fish Intake | 1329 | More than 1/wk: 3, 4, 5 | Yes/No |
| Non-Oily Fish Intake | 1339 | More than 1/wk: 3, 4, 5 | Yes/No |
| Smoking | 20116 | Yes: 2 | Yes/No |
| Alcohol | 20117 | Current: 2 | Yes/No |
| Prevalent Obesity | 130792 | Any < baseline date | Yes/No |
| Prevalent Hypertension | 131286, 131295 | Any < baseline date | Yes/No |
| Prevalent Diabetes | 130706, 130708, 130710, 130712, 130714 | Any < baseline date | Yes/No |
| Prevalent COPD | 131492 | Any < baseline date | Yes/No |
| Prevalent Renal Disease | 132032 | Any < baseline date | Yes/No |
| Prevalent MI | 131298, 131300, 131302, 131304,  131306 | Any < baseline date | Yes/No |
| Prevalent HF | 131354, 131288, 131292 | Any < baseline date | Yes/No |
| Hypertension Medication | 6153, 6177 | Yes: 2 | Yes/No |
| Hypertension Medication | 20003 | 1140860180, 1140862944, 1140863392, 1140865900, 1140867856, 1140868282, 1140868902, 1140869410, 1140869452, 1140869998, 1140871052, 1140872064, 1140872982, 1140874524, 1140874866, 1140875452, 1140876354, 1140876806, 1140877826, 1140878350, 1140878420, 1140879418, 1140879696, 1140879698, 1140881320, 1140881894, 1140882088, 1140882090, 1140882092, 1140882980, 1140883158, 1140888578, 1140888666, 1140888768, 1140909724, 1140909812, 1140910504, 1140910658, 1140910660, 1140910706, 1140910830, 1140916342, 1141145658, 1141146606, 1141150898, 1141171566, 1141175204, 1141176732, 1141178858, 1141179712, 1141180444, 1141180514, 1141180936, 1141181616, 1141181708, 1141188738, 1141195044, 2038459704 |  |
| Cholesterol Medication | 20003 | 1140861958, 1140873350, 1140873570, 1140874030, 1140874266, 1140874360, 1140880388, 1140880390, 1140882794, 1140882806, 1140882844, 1140882938, 1140883060, 1140888594, 1140888648, 1140910632, 1140910654, 1141146234, 1141157400, 1141192410 | Yes/No |
| Diabetes Medication | 20003 | 1140883066, 1140884600, 1141153254, 1141171646, 1141177600, 1141189090 | Yes/No |

**Supplemental Table 3. Using NMR-derived DHA and NMR Non-DHA Omega3 to predict GC-derived RBC or Plasma omega-3 levels**

| **Predicted^1^ Lipid Pool** | **Predicted GC FA metric** | **NMR DHA coefficient** | **NMR Non-DHA Omega-3 coefficient** | **Intercept** | **Model R^2^** |
| --- | --- | --- | --- | --- | --- |
| RBC | EPA+DHA | 2.629 | 0.4673 | -0.1014 | 67% |
|  | EPA | 0.901 | 0.227 | -1.253 | 64% |
|  | DHA | 1.728 | 0.240 | 1.150 | 56% |
|  | DPA | 0.450 | 0.155 | 1.634 | 34% |
| Plasma | EPA+DHA | 2.344 | 0.346 | -2.080 | 83% |
|  | EPA | 1.087 | 0.267 | -1.600 | 71% |
|  | DHA | 1.257 | 0.079 | -0.480 | 78% |
|  | DPA | 0.110 | 0.051 | 0.245 | 43% |

1. To predict a GC-based FA value, use the NMR DHA value for an individual, along with that same person’s NMR Non-DHA. For example, if an individual has an NMR DHA level of 2%, and a non-DHA Omega-3 level of 1%, the predicted RBC EPA+DHA value for that individual is equal to 2*2.629 + 1*0.4673 – 0.1014 = 5.62%.

**Supplemental Table 4 – Distribution of Variables Used in Zhang Re-evaluation.**

|  | **Overall** | **Non-Fish oil** | **Fish Oil** |
| --- | --- | --- | --- |
| Age 65+ | 87109 | 50814 | 36295 |
| White | 430666 | 293743 | 136923 |
| Male | 210365 | 147477 | 62888 |
| Townsend | -2.80258 | -1.25068 | -1.5519 |
| Oily Fish < 1 times/ week*** | 200955 | 149182 | 51773 |
| Non-oily fish < 1 times/week*** | 153293 | 111165 | 42128 |
| Current Smoking | 47809 | 36078 | 11731 |
| Current Drinking Alcohol | 419824 | 286020 | 133804 |
| Obesity | 11898 | 8508 | 3390 |
| Hypertension | 120410 | 80304 | 40106 |
| Diabetes mellitus | 22890 | 16228 | 6662 |
| COPD | 8273 | 5768 | 2505 |
| CRF | 5191 | 3565 | 1626 |
| CHD | 22779 | 15309 | 7470 |
| Heart failure | 1749 | 1232 | 517 |
| Antihypertensives | 93213 | 61665 | 31548 |
| Statins | 69220 | 45360 | 23860 |
| Antidiabetics | 15341 | 11097 | 4244 |
| Genetic Risk Score: |  |  |  |
| Low | 113642 | 77614 | 36028 |
| Intermediate | 227284 | 155409 | 71875 |
| High | 113642 | 77809 | 35833 |

**Supplemental Table 5: Absolute Risk for Atrial Fibrillation in Studies included in the meta-analysis by Jai et al.^2^ (with the addition of Miyauchi et al.^3^)**

| **All Studies** | N | | Events | | Absolute Risk | | |  |
| --- | --- | --- | --- | --- | --- | --- | --- | --- |
|  | Placebo | Active | Placebo | Active | Placebo | Active | Difference | Relative Risk |
| VITAL^4^ | 12577 | 12542 | 431 | 469 | 3.43% | 3.74% | 0.31% | 9% |
| ASCEND^5^* | 7740 | 7740 | 135 | 166 | 1.74% | 2.14% | 0.40% | 23% |
| STRENGTH^6^ | 6539 | 6539 | 86 | 144 | 1.32% | 2.20% | 0.89% | 67% |
| RP^7^ | 6266 | 6239 | 92 | 113 | 1.47% | 1.81% | 0.34% | 23% |
| REDUCE IT^8^ | 4090 | 4089 | 159 | 215 | 3.89% | 5.26% | 1.37% | 35% |
| DO-HEALTH^9^ | 1084 | 1073 | 67 | 78 | 6.18% | 7.27% | 1.09% | 18% |
| GISSI HF^10^ | 2914 | 2912 | 408 | 444 | 14.00% | 15.25% | 1.25% | 9% |
| OMEMI^11^ | 372 | 387 | 15 | 28 | 4.03% | 7.24% | 3.20% | 79% |
| RESPECT-EPA^3^ | 1235 | 1225 | 20 | 38 | 1.62% | 3.10% | 1.48% | 92% |
| Weighted Avg | 42817 | 42746 | 1413 | 1695 | **3.30%** | **3.97%** | **0.67%** | **20%** |
| **Low Dose (<1 g/d)** | N | | Events | | Absolute Risk | | |  |
|  | Placebo | Active | Placebo | Active | Placebo | Active | Difference | Relative Risk |
| VITAL | 12577 | 12542 | 431 | 469 | 3.43% | 3.74% | 0.31% | 9% |
| DO-HEALTH | 1084 | 1073 | 67 | 78 | 6.18% | 7.27% | 1.09% | 18% |
| ASCEND* | 7740 | 7740 | 135 | 166 | 7.60% | 7.70% | 0.10% | 1% |
| RP | 6266 | 6239 | 92 | 113 | 1.47% | 1.81% | 0.34% | 23% |
| GISSI HF | 2914 | 2912 | 408 | 444 | 14.00% | 15.25% | 1.25% | 9% |
| Weighted Avg | 30581 | 30506 | 1133 | 1270 | **3.70%** | **4.16%** | **0.46%** | **12%** |
| **High Dose (1.8 to 4 g/d)** | N | | Events | | Absolute Risk | | |  |
|  | Placebo | Active | Placebo | Active | Placebo | Active | Difference | Relative Risk |
| STRENGTH | 6539 | 6539 | 86 | 144 | 1.32% | 2.20% | 0.89% | 67% |
| REDUCE IT | 4090 | 4089 | 159 | 215 | 3.89% | 5.26% | 1.37% | 35% |
| OMEMI | 372 | 387 | 15 | 28 | 4.03% | 7.24% | 3.20% | 79% |
| RESPECT-EPA | 1235 | 1225 | 20 | 38 | 1.62% | 3.10% | 1.48% | 92% |
| Weighted Avg | 12236 | 12240 | 280 | 425 | **2.29%** | **3.47%** | **1.18%** | **52%** |

* EMR Data from ASCEND found no difference in AF events between the active and placebo groups^5^. When the EMR data are substituted for the originally published ASCEND results, the absolute risk difference in this study was 0.1% and the relative risk difference was 1%, resulting in absolute and relative risks for the low dose studies of 0.39% and 7%, and for the overall analysis, 0.61% and 14%, respectively.

1. Zhang J, Cai A, Chen G, et al. Habitual fish oil supplementation and the risk of incident atrial fibrillation: findings from a large prospective longitudinal cohort study. *European journal of preventive cardiology* 2022;29(14):1911-20. doi: 10.1093/eurjpc/zwac192 [published Online First: 2022/09/02]

2. Jia X, Gao F, Pickett JK, et al. Association Between Omega-3 Fatty Acid Treatment and Atrial Fibrillation in Cardiovascular Outcome Trials: A Systematic Review and Meta-Analysis. *Cardiovascular Drugs and Therapy* 2021;35(4):793-800. doi: 10.1007/s10557-021-07204-z

3. Miyauchi K, Iwata H, Nishizaki Y, et al. Randomized Trial for Evaluation in Secondary Prevention Efficacy of Combination Therapy–Statin and Eicosapentaenoic Acid (RESPECT-EPA). *Circulation* 2024;150(6):425-34. doi: 10.1161/CIRCULATIONAHA.123.065520

4. Albert CM, Cook NR, Pester J, et al. Effect of Marine Omega-3 Fatty Acid and Vitamin D Supplementation on Incident Atrial Fibrillation: A Randomized Clinical Trial. *Jama* 2021;325(11):1061-73. doi: 10.1001/jama.2021.1489 [published Online First: 2021/03/17]

5. Parish S, Mafham M, Offer A, et al. Effects of Omega-3 Fatty Acid Supplements on Arrhythmias. *Circulation* 2020;141(4):331-33. doi: doi:10.1161/CIRCULATIONAHA.119.044165

6. Nicholls SJ, Lincoff AM, Garcia M, et al. Effect of High-Dose Omega-3 Fatty Acids vs Corn Oil on Major Adverse Cardiovascular Events in Patients at High Cardiovascular Risk: The STRENGTH Randomized Clinical Trial. *JAMA* 2020;324:2268-80. doi: 10.1001/jama.2020.22258

7. Roncaglioni MC, Tombesi M, Avanzini F, et al. n-3 fatty acids in patients with multiple cardiovascular risk factors. *The New England journal of medicine* 2013;368(19):1800-8. doi: 10.1056/NEJMoa1205409 [published Online First: 2013/05/10]

8. Bhatt DL, Steg PG, Miller M, et al. Cardiovascular Risk Reduction with Icosapent Ethyl for Hypertriglyceridemia. *The New England journal of medicine* 2019;380(1):11-22. doi: 10.1056/NEJMoa1812792 [published Online First: 2018/11/13]

9. Bischoff-Ferrari HA, Vellas B, Rizzoli R, et al. Effect of Vitamin D Supplementation, Omega-3 Fatty Acid Supplementation, or a Strength-Training Exercise Program on Clinical Outcomes in Older Adults: The DO-HEALTH Randomized Clinical Trial. *Jama* 2020;324(18):1855-68. doi: 10.1001/jama.2020.16909 [published Online First: 2020/11/11]

10. Investigators G-H. Effect of n-3 polyunsaturated fatty acids in patients with chronic heart failure (the GISSI-HF trial): a randomised, double-blind, placebo-controlled trial. *Lancet* 2008;372:1223-30.

11. Myhre PL, Kalstad AA, Tveit SH, et al. Changes in eicosapentaenoic acid and docosahexaenoic acid and risk of cardiovascular events and atrial fibrillation: A secondary analysis of the OMEMI trial. *J Intern Med* 2022;291(5):637-47. doi: 10.1111/joim.13442 [published Online First: 2022/01/05]
